# Methodological Considerations for Linking Household and Healthcare Provider Data for Estimating Effective Coverage: A Systematic Review

**DOI:** 10.1101/2020.09.28.20203216

**Authors:** Emily D. Carter, Hannah H. Leslie, Tanya Marchant, Agbessi Amouzou, Melinda Munos

## Abstract

Effective coverage measures assess the proportion of a population that receive a health intervention with sufficient quality to achieve health benefit. Linking population-based surveys and health facility data is a promising means of generating effective coverage estimates, however, little guidance exists on methodological considerations for these analyses.

We conducted a systematic review to assess existing knowledge related to 1) the suitability of data used in linking analyses, 2) the implications of the design of existing data sources commonly used in linking analyses, and 3) the impact of choice of method for combining datasets to obtain linked coverage estimates. The primary search was completed in Medline, with additional reviews of select sources.

Of 3192 papers reviewed, 62 publications addressed issues related to linking household and provider datasets. Limited data suggest household surveys can be used to identify sources of care, but their validity in estimating a denominator of intervention need was variable. Methods for collecting provider data and constructing quality indices were variable and presented limitations. There was little empirical data supporting an association between structural, process, and outcome quality. Few studies addressed the influence of the design of common data sources on linking analyses, including imprecise household GIS data, provider sampling frame and sampling design, and estimate stability. There was a lack of concrete evidence around the impact of these factors on linked effective coverage estimates. The most consistent evidence suggested under certain conditions, combining data sets based on geographical proximity (ecological linking) produced similar estimates to linking based on the specific provider utilized (exact-match linking).

Linking household and healthcare provider can leverage existing data sources to generate more informative estimates of intervention coverage and care. However, there is need for additional research to develop evidence-based, standardized best practices for these analyses.

**What is already known?:**

- Linking population-based and provider data is a means of generating effective coverage estimates, however little guidance exists on methodological considerations for linking these data sources

**What are the new findings?:**

- 62 publications address issues related to the 1) the suitability of data used in linking analyses, 2) the implications of the design of existing data sources commonly used in linking analyses, and 3) the impact of choice of method for combining datasets to obtain linked coverage estimates
- There was variable and limited evidence on the suitability of data household and provider data, particularly collecting and constructing indicators of provider quality, and on the implications of the design of existing data sources
- The most consistent evidence suggested under certain conditions, combining data sets based of geographical proximity or administrative unit produced similar estimates to linking based on the specific provider utilized

**What do the new findings imply?:**

- There is need for additional research to develop evidence-based, standardized best practices for linked analyses of health system and population data

## BACKGROUND

There is growing demand for tracking progress towards the sustainable development goals (SDGs) through effective coverage estimates [1,2]. Effective coverage measures assess not only the proportion of individuals in need of an intervention who receive it, but also the content and quality of services received with an aim to estimate the proportion of individuals receiving the health benefit of an intervention [2]. Numerous publications have estimated effective coverage [3] using a range of methods and measures to define intervention need, receipt, and quality.

Linking household and health provider data is a promising means of generating effective coverage estimates that provide population-based estimates and incorporate data on service quality from health facilities. Data from household surveys can provide a population-based estimate of intervention need and care-seeking for services, such as the proportion of women with a recent live birth who delivered in a health facility. However, a number of maternal, newborn, and child health interventions [4] cannot be accurately measured through household surveys due to reporting errors and biases by respondents (e.g., the proportion of women who received a uterotonic during delivery). Health provider assessments yield information on provider quality, including available infrastructure, commodities, equipment, human resources, and potentially provision of care. Provider data do not capture need for care in the population, care-seeking behavior, or the experience of individuals who do not access the formal health system. Linking these two data sources can provide a more complete picture of population access to and coverage of high-quality health services, for example the proportion of women who delivered at a health facility with sufficient structural resources and competence to provide appropriate labor and delivery care.

There are many approaches for combining household and provider datasets [5]. The results depend on the choice of data and of methods for combining datasets. However, very limited guidance exists to guide decision making. We conducted a systematic review to understand the current evidence base for effective coverage linking methods and identify needs for further research.

## METHODS

We searched for papers addressing methods or assumptions regarding: 1) the suitability of household and provider ^1^ data used in linking analyses, 2) the implications of the design of existing household (DHS and MICS) and provider (SPA and SARA) data sources commonly used in linking analyses, and 3) the impact of choice of method for combining datasets to obtain linked coverage estimates.

Our primary search was conducted in Medline. The search was limited to papers published between January 1, 2000 and June 7, 2019 that included terms related to 1) effective coverage, benchmarking, system dynamics, or universal health coverage metrics, or 2) structural, process, and/or health outcome quality, 3) linking analyses using terms adapted from Do and colleagues [5], 4) validity of self-report health indicators, and 5) spatial methods for measuring utilization or distance to care. A full list of Medline search terms and PRISMA checklist are presented in Supplementary File 1 and 2, respectively. The search was conducted using English-language terms; however, publications in English, Spanish, and French were reviewed if captured in the search. Additionally, we conducted searches using these criteria health workers in Population Health Metrics (which was not fully indexed in Medline at the time of our search), the Carolina Population Health Center, and DHS publications. In a second step, we hand-searched the references of a systematic review by Do and colleagues on linking household and facility data to estimate coverage of reproductive, maternal, newborn, and child health services [5], and a review by Amouzou and colleagues of effective coverage analyses [3]. We also hand-searched the references, citing works, and journal- or database interface-generated related publications of all articles that passed the title and abstract review. Particularly relevant papers published after the formal search date were included in the review if captured through these snowball review mechanisms.

Publications were reviewed for relevant analyses or commentary related to linking methodologies. Articles were included if they explicitly evaluated or compared assumptions used in linking approaches for at least one of the areas defined above. Title and abstract review were conducted simultaneously. Data extraction was completed by the first author (EC) and included the title, author, year of publication, country or countries included in analysis, data source, and specific analyses or findings relevant to linking loosely categorized by topic areas.

## RESULTS

The Medline search produced 3055 publications, along with 97 from the Carolina Population Center, 4 from Population Health Metrics, 12 DHS publications, 35 papers included in the review by Amouzou and colleagues and 49 papers included in the review by Do and colleagues meeting the publication date restrictions. After removing duplicates, 3192 publications were included in the title and abstract review and 218 were included in the full text review. Of those papers included in the full text review, 48 publications addressed a methodological concern related to linking household and provider data and were included in the final review. Fourteen additional publications were identified through the snowball review of references and related works (Fig 1 – PRISMA flow diagram). In total 62 publications addressed a methodological concern, including the suitability of household (n=13) and provider data (n=32) for use in linking analyses, concerns related to the design of existing household (n=6) and provider (n=4) data sources, and methods for combining household and facility data (n=13). A list of publications included in the review and a summary of their contributions to the review are provided in Table 1.

**Table 1.** Summary of publications included in the review and contribution to the literature

| AUTHOR | YEAR | COUNTRY | KEY METHOD CONTRIBUTION |
| --- | --- | --- | --- |
| **SUITABILITY OF HOUSEHOLD AND PROVIDER DATA FOR LINKING ANALYSES** |  |  |  |
| • **Household data needed for linked estimates** |  |  |  |
| Ayede [7] | 2018 | Nigeria | • Accuracy of maternal report of pneumonia symptoms measured through household survey |
| Blanc [16] | 2016 | Mexico | • Accuracy of maternal report of delivery / immediate PNC attendant measured through household survey |
| Blanc [17] | 2016 | Kenya | • Accuracy of maternal report of delivery / immediate PNC attendant measured through household survey |
| Carter [14] | 2018 | Zambia | • Accuracy of maternal report of care-seeking for child illness measured through household survey |
| Chang [13] | 2018 | Nepal | • Accuracy of maternal report of birthweight and preterm birth measured through household survey |
| D'Acremont [9] | 2010 | SSA | • Reduced proportion of fever cases that are malaria |
| Hazir [8] | 2013 | Pakistan & Bangladesh | • Accuracy of maternal report of pneumonia symptoms measured through household survey |
| Keenan [11] | 2017 | US | • Accuracy of maternal recall of birth complications |
| Shengelia [6] | 2005 | - | • Effect of true versus perceived intervention need on effective coverage estimation |
| Stanton [15] | 2013 | Mozambique | • Accuracy of maternal report of place of delivery measured through household surveys |
| Walker [10] | 2013 | - | • Issues with measurement of child diarrhea through household surveys |
| Wang [18] | 2018 | Multiple Regions | • Issues with provider categories and alignment between DHS and SPA surveys |
| Zimmerman [12] | 2019 | Ethiopia | • Reliability of maternal report of maternal and newborn birth complications |
| **SUITABILITY OF HOUSEHOLD AND PROVIDER DATA FOR LINKING ANALYSES** |  |  |  |
| • **Provider data needed for linked estimates** |  |  |  |
| Akachi [49] | 2017 | - | • Need for global benchmarks for quality<br>• Lack on data linking quality with health outcomes |
| Carter [31] | 2018 | Zambia | • Quality score for child health effective coverage |
| Davis [30] | 2006 | High-income countries | • Agreement between provider self-assessment and observed quality |
| Diamond-Smith [23] | 2016 | Kenya & Namibia | • Association between maternal perception of care and measured structural and process quality |
| Fisseha [43] | 2017 | Ethiopia | • Internal consistency of structural and process quality indicator |
| Gabrysch [40] | 2011 | Zambia | • Quality score for labor and delivery effective coverage |
| Hoogenboom [25] | 2015 | Thai-Myanmar Border | • Agreement between facility records and observed care |
| Hrisos [28] | 2009 | High-income countries | • Systematic review of agreement between observed quality of care and provider self-report, patient-report, and/or chart review |
| Jackson [45] | 2015 | Tanzania | • PCA to reduce quality index |
| Kanyangarara [35] | 2017 | SSA | • Quality score for ANC effective coverage |
| Kruk [47] | 2017 | SSA | • Association between structural and process quality |
| Larson [24] | 2014 | Tanzania | • Association between maternal perception of care and service availability and respect<br>• Vignettes for measuring quality |
| Leegwater [51] | 2015 | - | • Association between UHC index and infant mortality and life expectancy at national level |
| Leslie [50] | 2016 | Malawi | • Association between quality of delivery care and neonatal mortality |
| Leslie [38] | 2017 | SSA | • Quality score for ANC, labor and delivery, sick child, and family planning effective coverage |
|  |  |  | • Association between structural and process quality |
| Leslie [46] | 2018 | Multiple Regions | • Performance of approaches for generating service readiness indices |
| Lozano [41] | 2006 | Mexico | • UHC index using weighted vs simple average of indicators |
| Mallick [44] | 2017 | Haiti, Malawi & Tanzania | • Comparison of measures of family planning quality |
| Marchant [33] | 2015 | Ethiopia, Nigeria, & India | • Measurement of quality using "last delivery module" |
| Mboya [34] | 2016 | Tanzania | • mHealth tool to measure quality |
| MCSP [22] | 2018 | Multiple Regions | • Availability and quality of data captured through HMIS |
| Munos [37] | 2018 | Cote D'Ivoire | • Quality score for ANC, labor and delivery, PNC, and child health effective coverage |
| Nesbitt [26] | 2013 | Ghana | • Quality score for labor and delivery and PNC effective coverage |
| Nguhiu [36] | 2017 | Kenya | • Quality score for ANC, labor and delivery, sick child, and family planning effective coverage |
| Nickerson [20] | 2015 | Multiple Regions | • Comparison of data collected through health facility assessments |
| Osen [27] | 2011 | Ghana | • Agreement between provider reported and observed surgical service quality |
| Peabody [29] | 2000 | US | • Agreement between vignettes, chart abstraction, and simulated client measures |
| Sheffel [21] | 2018 | Multiple Regions | • Summary of quality data collected through SPA and SPA |
| Sheffel [42] | 2018 | Haiti, Malawi, Tanzania | • Association between structural and process quality |
| Willey [39] | 2018 | Uganda | • Quality score for labor and delivery and newborn care effective coverage |
| Wilunda [32] | 2015 | Uganda | • Quality score for maternal and neonatal care effective coverage |
| Zurovac [48] | 2015 | Vanuatu | • Poor association between structural quality and clinical care in fever management |
| **IMPLICATIONS OF DESIGN OF EXISTING HOUSEHOLD AND PROVIDER DATA SOURCES COMMONLY USED IN LINKING ANALYSES** |  |  |  |
| • Household data |  |  |  |
| Bliss [54] | 2012 | USA | • Comparison of distance using centroid versus true location |
| Healy [56] | 2012 | Canada & UK | • Comparison of distance using centroids of varying areal groupings |
| Jones [55] | 2010 | US | • Comparison of distance using zip-code centroid versus true household location |
| Nesbitt [57] | 2014 | Ghana | • Comparison of straight-line distance, network distance, raster and network-based travel time distance measures using village versus compound centroid |
| Perez-Heydrch [52] | 2013 | - | • Effect of DHS cluster displacement on distance measures |
| Skiles [58] | 2013 | Rwanda | • Effect of DHS cluster displacement on estimates of service environment |
| **IMPLICATIONS OF DESIGN OF EXISTING HOUSEHOLD AND PROVIDER DATA SOURCES COMMONLY USED IN LINKING ANALYSES** |  |  |  |
| • Provider data |  |  |  |
| Carter [31] | 2018 | Zambia | • Effect of excluding non-facility providers from sampling frame on effective coverage estimates |
| Munos [37] | 2018 | Cote d'Ivoire | • Effect of excluding non-facility providers from sampling frame on effective coverage estimates |
| Skiles [58] | 2013 | Rwanda | • Effect of facility sampling on estimates of service environment |
| Turner [59] | 2001 | - | • Limitations of SPA sampling design<br>• Approach for joint sampling of households and facilities for linking analyses |
| **IMPLICATIONS OF DESIGN OF EXISTING HOUSEHOLD AND PROVIDER DATA SOURCES COMMONLY USED IN LINKING ANALYSES** |  |  |  |
| • Survey timing |  |  |  |
| Baker [60] | 2005 | Uganda & Tanzania | Stability of facility diagnostic capacity over time |
| Marchant [61] | 2008 | Tanzania | Stability of IPTp stocks |
| Wang [63] | 2011 | Multiple Regions | Stability of maternal health care-seeking behaviors measured through household survey over time |
| Willey [39] | 2018 | Uganda | Stability of facility infrastructure indicators for labor, delivery, and newborn care |
| Winter [62] | 2015 | Multiple Regions | Stability of care-seeking for child illness behaviors measured through household survey over time |
| **IMPACT OF CHOICE OF METHOD FOR COMBINING HOUSEHOLD AND PROVIDER DATA** |  |  |  |
| Comparison of exact-match and ecological linking methods for estimating effective coverage |  |  |  |
| Carter [31] | 2018 | Zambia | Comparison of exact-match and ecological linking methods in estimating effective coverage in sick child care |
| Munos [37] | 2018 | Cote d'Ivoire | Comparison of exact-match and ecological linking methods in estimating effective coverage in ANC, labor and delivery, PNC, and sick child care |
| Willey [39] | 2017 | Uganda | Comparison of exact-match and ecological linking methods in estimating effective coverage in ANC, labor and delivery, PNC, and sick child care |
| **IMPACT OF CHOICE OF METHOD FOR COMBINING HOUSEHOLD AND PROVIDER DATA** |  |  |  |
| Performance of measures of geographic proximity for ecological linking |  |  |  |
| Carter [31] | 2018 | Zambia | Comparison of true-source of care for child illness to straight-line and road distance measures |
| Delamater [66] | 2019 | US | Comparison of FCA, simple distance, and Huff distance measure against true utilization patterns |
| Gething [68] | 2004 | Kenya | Comparison of Theissen boundaries and true utilization patterns |
| Munos [37] | 2018 | Cote d'Ivoire | Comparison of true-source of care for ANC, labor and delivery, PNC, and child illness to straight-line and road distance measures |
| Noor [64] | 2006 | Kenya | Comparison of true-source of care for child fever to closest by Euclidian and road distance |
| Tanser [69] | 2001 | South Africa | Comparison of Theissen boundaries and true utilization patterns |
| Tanser [65] | 2006 | South Africa | Comparison of typical source of care to closest by travel time |
| Tsoka [67] | 2004 | South Africa | Comparison of Theissen boundaries and true utilization patterns |
| **IMPACT OF CHOICE OF METHOD FOR COMBINING HOUSEHOLD AND PROVIDER DATA** |  |  |  |
| Statistical challenges |  |  |  |
| Wang [18] | 2018 | Multiple Regions | Use of Delta method for estimating effective coverage variance |
| Willey [39] | 2018 | Uganda | Use of Delta method for estimating effective coverage variance |

**Figure 1.**
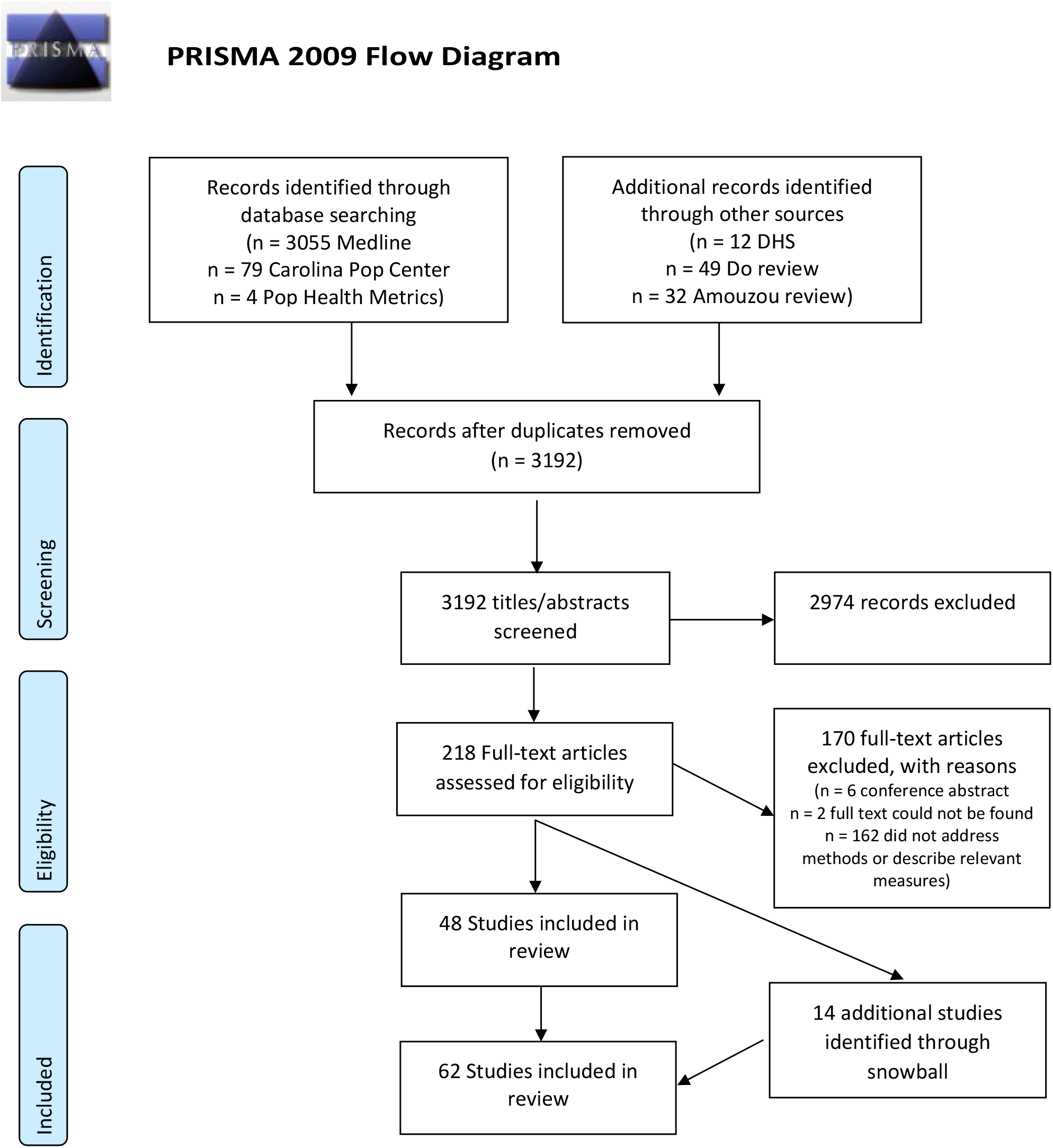
PRISMA Flow Diagram.

### Suitability of household and provider data for linking analyses

#### Suitability of household data needed for linked estimates

In effective coverage linking analyses, household surveys can be used to estimate the population in need of healthcare, as well as care-seeking behavior. Household surveys must produce valid estimates of these parameters and provide care-seeking data that can be linked to provider assessments. This review identified papers discussing issues in defining intervention need (n=8) and care-seeking (n=5) that should guide selection of indicators for linking.

##### Intervention need

Estimation of intervention need may require solely population demographics such as age (e.g., for prevention and health promotion interventions) or may require defining specific illnesses or conditions The latter is more subject to reporting bias [6]. Multiple studies have shown poor association or biases between maternally-reported symptoms and clinical pneumonia [7,8], malaria [9], and diarrhea [10] in children under five. A handful of studies (n=3) showed maternal report of both maternal and newborn birth complications is variable [11–13]. A simulation by Shengelia and colleagues demonstrated the effect of the divergence of true from perceived intervention need on effective coverage estimates. The authors propose estimating the posterior probability of disease based on responses to symptomatic questions using a Bayesian model to measure disease presence on a probabilistic scale [6]. However, there has been no work on how to integrate these adjusted estimates into effective coverage estimates.

##### Care-seeking behavior

Four studies addressed the accuracy of respondent report on seeking care. Mothers in Zambia and Mozambique were able to accurately report on the type of health provider where they sought sick child [14] and delivery care [15], respectively. However, studies in two countries suggested women cannot report on the type of healthworker who attended to them during labor and delivery and immediate post-natal care [16,17]. Wang and colleagues note that provider categories are not standardized between population surveys and health system assessments, with population surveys often including vague or overly broad categories that do not directly match SPA/SARA categories and require harmonization [18].

##### Suitability of healthcare quality data needed for linked estimates

Provider assessments present data on service content and quality for effective coverage linking analyses. However, the measurement, construction, and interpretation of provider quality measures are highly variable and may significantly alter effective coverage estimates. This paper does not present an exhaustive review of healthcare quality measures or the association between levels of quality. A comprehensive summary of quality of care concepts and measurement approaches, along with their relative strengths and limitations, was presented by Hanefeld and colleagues [19]. Publications of particular relevance to linking analyses are noted here, with an emphasis on national provider survey data as the most common source of provider data for linking analyses.

##### Methods used in assessing provider quality

A review by Nickerson and colleagues found significant variability in the data collected and methods used in health facility assessment tools in LMICs [20]. While SPA and SARA data are the most widely used sources of data on health service delivery in LMICs, one paper noted that these surveys focused primarily on structural quality with less data on provision and experience of care [21]. The lack of process quality data is in part related to the reliance on direct observation of clinical care – a time- and resource-intensive method – to collect these data. None of the studies included in the review used HMIS data to generate linked coverage estimates. A desk review by MCSP found that data collected through HMIS was variable across countries, data recorded within registers often was not transmitted through the system, and only a limited number of indicators collected were related to the provision of health services [22].

Eight publications assessed alternatives to direct observation of clinical care for collecting process quality data. Two studies found variable association between process quality and maternal perceptions of the quality of care received [23,24]. Agreement between observed care and health records or provider report was also variable [25–27]. A review by Hrisos and colleagues found few studies to support use of patient report, provider self-report, or record review as proxy measures of clinical care quality [28]. In the US, vignettes performed better than chart abstraction for estimating quality [29]. Another review found providers were unable to accurately assess their own performance, with the worst accuracy among the least skilled providers [30]. Five other publications used alternative methods for measuring process quality, including use of vignettes [24,31], register review [24,32], most recent delivery interview [33], and an mHealth tool [34], but did not assess their performance against other measurement methods.

##### Content of provider quality indices

Most linking papers estimating effective coverage included in this review (n=10) characterized provider quality using structural measures of quality, with or without measures of process quality. Various approaches were used to select items for inclusion in these measures. Measures of structural and process quality were derived from either national or international guidance on minimum service availability and required commodities, equipment, infrastructure, training, or actions. Measures used by effective coverage analyses included SPA or SARA structural indicators [31,35,36] and/or clinical observations [36–38], EmOC functions [26,32,39,40], and provider recall of actions during their last delivery [33,39].

##### Construction of provider quality indices

In addition to the range of variables used in provider measures, there was no consensus on the approach to use to select and combine variables to generate quality indices. The reviewed publications used a variety of approaches to construct indices including weighted indices [38], simple averages across all indicators or domains [31,37,39], and categorization using set thresholds or relative categories [24,35,40]. Seven publications presented data on the performance of different measurement modes and summary approaches. Two studies found the method of selecting and combining quality indicators had little effect on overall effective coverage estimates [41,42]. However, two other studies found inconsistency in the rankings of health facilities when using different index methods [43,44]. Two studies using PCA to create SPA health service indices found the reduced indices explained only a limited amount of the variance across indicators [44,45]. An analysis of SPA data in ten countries found indices empirically-derived through machine learning captured a large proportion of the service readiness data in the full SPA index, however the selected set of indicators varied across countries, and an index generated through expert review captured very little of the data from the full index [46]. Two studies found that few facilities could meet all requirements when applying a threshold, limiting the utility of the approach [40,43].

##### Performance of provider measures

Despite the common usage of SPA and SARA data-derived structural and process quality measures, the review found limited data explicitly assessing the association of these measures with each other and health outcomes (n=7). Three studies, two incorporating data from multiple countries, found little association between structural quality and process quality [38,47,48]. However, an analysis of SPA data from three countries found a small but significant association between ANC facility structural and process quality and suggests structural quality can limit provider performance when basic infrastructure and commodities are unavailable [42]. Akachi and Kruk emphasized the limited number of studies showing process quality associated with health outcomes [49]. One study found a small association between an obstetric quality index and decreased neonatal mortality in Malawi [50] and another found a national UHC “heath service coverage” index correlated strongly with infant mortality rate and life expectancy [51].

### Implications of design of existing household and provider data sources commonly used in linking analyses

#### Issues related to household and cluster location data

The way in which common household surveys, particularly the DHS and MICS, collect and process location data may also impact the validity of some linked estimates. In many household datasets used for linked analyses, the precise location of individual households is often unknown. The DHS collects central point locations for clusters, rather than household locations, and displaces these points in publicly released datasets [52]. MICS often does not collect or make GIS data available [53]. Imprecision around household location may influence the accuracy of estimates generated by linking household and provider data based on geographic proximity.

##### Data on household location

The effect of using cluster central point locations (centroids) rather than individual household locations in linking analyses was not addressed by any publication identified in this review. However, four studies looked at the effect of using centroids of varying areal units versus household locations in distance analyses. Two studies found using US census tract [54] and zip-code [55] centroid locations produced little difference in measures of facility access compared to household location. A third study showed use of areal unit centroids resulted in misclassification of household access to health-related facilities, especially in less densely populated rural areas [56]. However, in rural Ghana, measures calculated from village centroids identified the same closest facility as measures from compound locations for over 85% of births [57].

##### Cluster displacement

Displacement of cluster central points might induce additional error in analyses based on geographic proximity. A DHS analytic report found that ignoring DHS displacement in analyses that used distance to a resource as a covariate resulted in increased bias and mean squared error (MSE). However, this will not affect linking by administrative unit because DHS has restricted displacement to within the representative sample administrative unit since 2009 [52]. A simulation analysis in Rwanda reported DHS cluster displacement produced less misclassification in level of access and relative service quality than healthcare provider sampling [58].

#### Issues related to provider sampling

Typical sampling designs for healthcare provider data also present issues for linking analyses. Both SPAs and SARAs are sampled independently of household surveys, thus, there may be no sampled facilities near household survey clusters [59]. SPA and SARA surveys typically collect data on a sample, rather than census, of public, private, and NGO health facilities and exclude non-facility providers, such as pharmacies or community health workers. In most settings, facilities are sampled and analyzed to be representative of all facilities within a managing authority, level, and/or geographic area, and the results of the provider assessment are not intended to represent the population utilizing health services [59]. For provider assessments conducting direct observations of clinical care, the number and type of interactions observed within each health facility is dependent on patient volume and chance.

##### Provider sampling frame

Two papers assessed the impact of excluding non-facility providers on linked effective coverage estimates. In Zambia and Cote d’Ivoire, CHWs offered a level of care for sick children similar to first-level public facilities. Excluding these providers reduced estimates of effective coverage in Zambia where CHWs were a significant source of skilled care in rural areas estimates [31], but had little effect in Cote d’Ivoire where they were an insignificant source of care [37]. In both studies, exclusion of pharmacies did not alter effective coverage estimates as they were an uncommon source of care, though they offered moderate structural quality [31,37].

##### Provider sampling design

Two publications addressed the impact of facility survey sampling designs. At the facility level, Skiles and colleagues’ analysis demonstrated that sampling facilities, rather than using a census, led to an underestimation of the adequacy of the health service environment and substantial misclassification error in relative service environment for individual clusters [58]. No studies addressed the suitability of SPA or SARA facility sampling strategies for generating stable quality estimates for use in linking analyses at a level below administrative unit used for the sampling approach.

A Measure Evaluation manual emphasized that data on provision of services (collected through observation of client-staff interactions) and experience of care (collected through client exit interviews) are a sub-sample of the overall survey and representative at the level the survey is sampled to be representative – not at the facility level [59]. This paper proposed multiple linked sampling approaches to capture geographically concordant household and provider data for linked analyses. While multiple studies included in this review used a census or sample of providers derived from a household sample, none implemented this approach at a national scale.

#### Issues related to timing of surveys used in linked coverage estimates

Both care-seeking behavior and provider quality are likely to vary over time, and both household and provider surveys are conducted infrequently in LMICs (∼3-5 years). Linked coverage estimates for RMNCH may cover a long timeframe as the reference period for care-seeking in household surveys varies from 2 weeks (sick child care) to 2-5 years (peripartum care). Population movement and quality improvement efforts at facilities further complicate associations with increasing time lags. The implications of linking household and provider indicators of different temporal periods is unclear.

##### Stability of provider indicators

No paper in this review specifically addressed the effect of provider indicator stability on linked effective coverage estimates. However, three linking papers presented data on the stability of some health facility indicators over time. EQUIP studies in Uganda and Tanzania found moderate variability in the availability of some maternal and newborn health commodities and services over a 2-3 year period [39,60,61].

##### Stability of household indicators

Care-seeking behavior, including overall rates of care and utilization of different sources of care, may also change over time. Analysis of care-seeking for child illness [62] and maternal healthcare [63] in multiple low- and middle-income countries over time showed high inconsistency in trends across countries. However, no identified studies addressed the consequences of this temporal variability within the context of linking analyses.

### Impact of choice of method for combining household and provider data

The approach for combining household and provider data can potentially have a significant impact on linked coverage estimates. Methods used to link data, including exact-match and various types of ecological linking, are defined in Table 2. Exact-match linking assigns provider information to individuals in the target population based on their specific source of care. This approach, while potentially subject to the reporting biases described previously, is considered the most precise approach for combining the two data sets in the absence of individual patient health records [5]. Without data on specific source of care, ecological linking approaches are designed to approximate care-seeking behavior or model healthcare access by linking the target population to sources of care based on geographic proximity or administrative catchment area, making assumptions about service access and use.

**Table 2.**
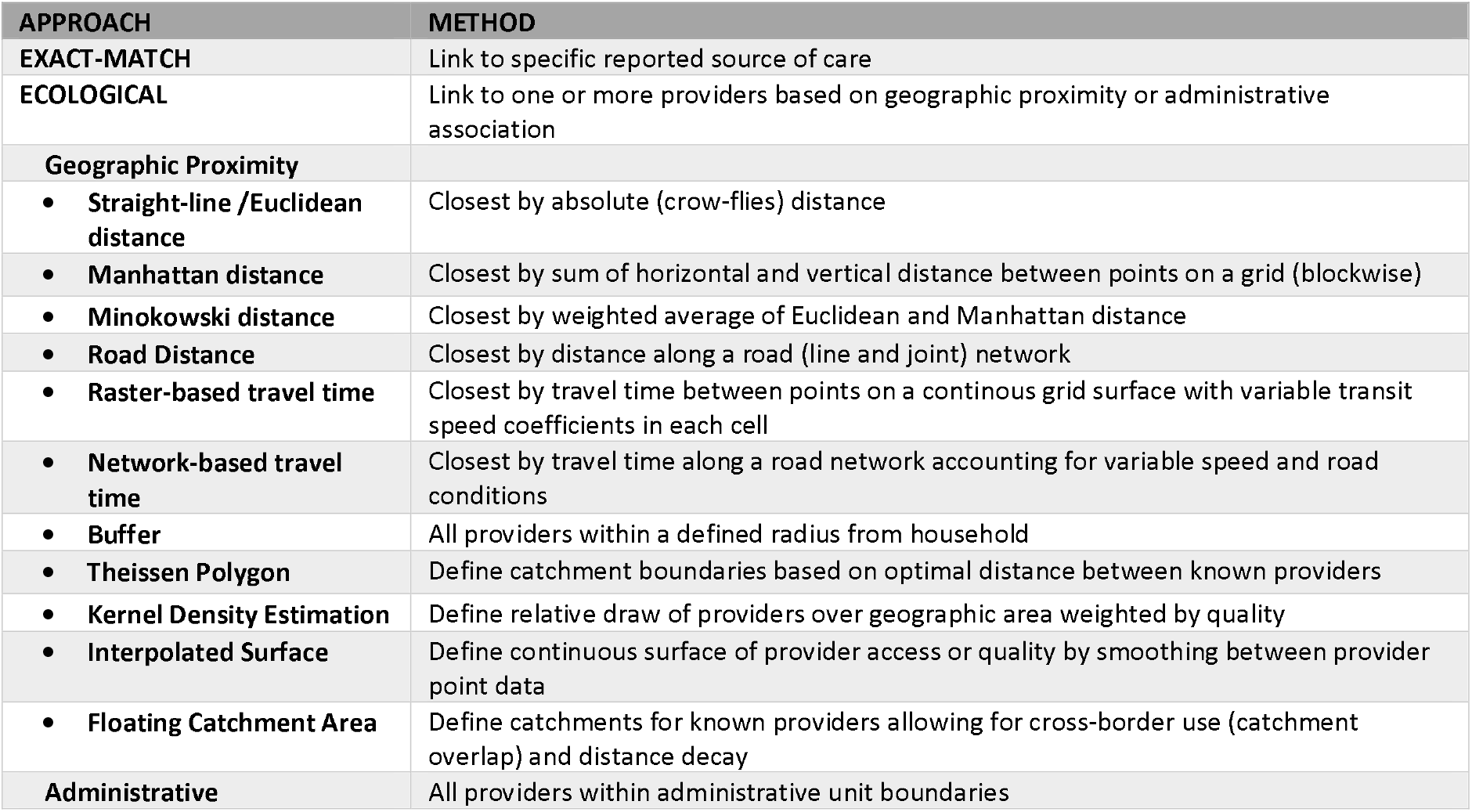
Table of linking approaches

#### Comparison of exact-match and ecological linking methods for estimating effective coverage

Three publications explicitly compared effective coverage estimates generated using exact-match and ecological linking methods (Table 3) [31,37,39]. Estimates generated using the exact-match linking approach were considered the gold-standard measure of effective coverage. All three publications found exact-match linked effective coverage estimates were similar to straight-line [31,37], travel time [31,37], 5-km buffer [31], 10-km buffer [37], and administrative unit [31,37,39] geolinked estimates for antenatal [37], labor and delivery [37,39], postnatal [37] and sick child [31,37] care when linking was restricted by the reported provider category (e.g., hospital, health center, community health worker). Distance-restricted linking approaches, such as linking to providers within a 5 km radius, produced inaccurate results if unlinked events were treated as no care [31]. Restriction of geographic linking to only providers within the reported category of care and/or weighting by providers’ relative patient volume improved agreement between the exact-match and ecological linking estimates [37,39]. All three studies also used provider data obtained from a census of health facilities, and therefore the findings may not be applicable when household data are linked to a sample survey of health facilities.

**Table 3.**
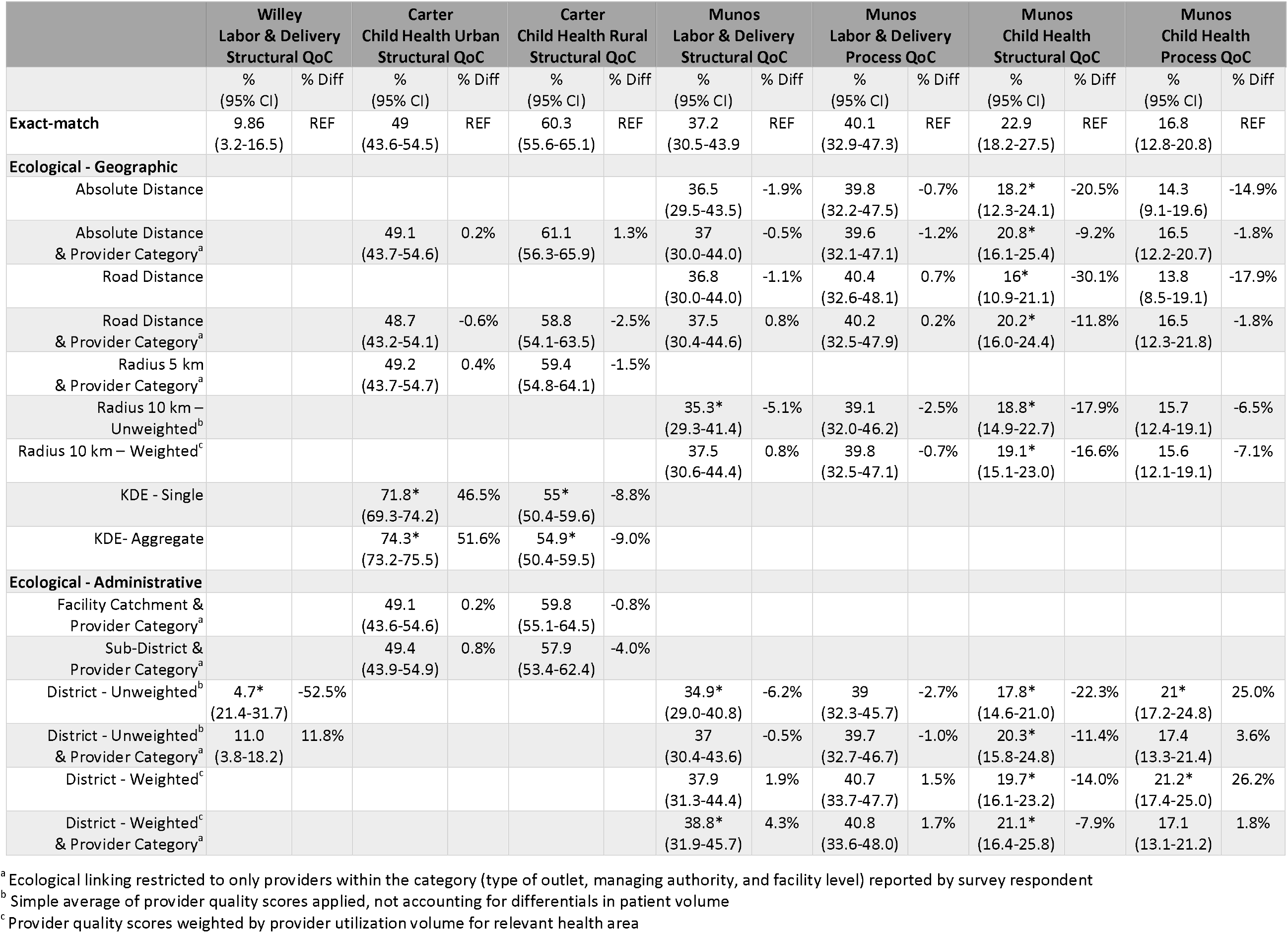
Exact versus ecological linking estimates for select indicators across studies

|  | Wiley<br>Labor & Delivery<br>Structural QoC |  | Carter<br>Child Health Urban<br>Structural QoC |  | Carter<br>Child Health Rural<br>Structural QoC |  | Munos<br>Labor & Delivery<br>Structural QoC |  | Munos<br>Labor & Delivery<br>Process QoC |  | Munos<br>Child Health<br>Structural QoC |  | Munos<br>Child Health<br>Process QoC |  |
| --- | --- | --- | --- | --- | --- | --- | --- | --- | --- | --- | --- | --- | --- | --- |
|  | %<br>(95% CI) | % Diff | %<br>(95% CI) | % Diff | %<br>(95% CI) | % Diff | %<br>(95% CI) | % Diff | %<br>(95% CI) | % Diff | %<br>(95% CI) | % Diff | %<br>(95% CI) | % Diff |
| **Exact-match** | 9.86<br>(3.2-16.5) | REF | 49<br>(43.6-54.5) | REF | 60.3<br>(55.6-65.1) | REF | 37.2<br>(30.5-43.9) | REF | 40.1<br>(32.9-47.3) | REF | 22.9<br>(18.2-27.5) | REF | 16.8<br>(12.8-20.8) | REF |
| **Ecological - Geographic** |  |  |  |  |  |  |  |  |  |  |  |  |  |  |
| Absolute Distance |  |  |  |  |  |  | 36.5<br>(29.5-43.5) | -1.9% | 39.8<br>(32.2-47.5) | -0.7% | 18.2*<br>(12.3-24.1) | -20.5% | 14.3<br>(9.1-19.6) | -14.9% |
| Absolute Distance<br>& Provider Category<sup>a</sup> |  |  | 49.1<br>(43.7-54.6) | 0.2% | 61.1<br>(56.3-65.9) | 1.3% | 37<br>(30.0-44.0) | -0.5% | 39.6<br>(32.1-47.1) | -1.2% | 20.8*<br>(16.1-25.4) | -9.2% | 16.5<br>(12.2-20.7) | -1.8% |
| Road Distance |  |  |  |  |  |  | 36.8<br>(30.0-44.0) | -1.1% | 40.4<br>(32.6-48.1) | 0.7% | 16*<br>(10.9-21.1) | -30.1% | 13.8<br>(8.5-19.1) | -17.9% |
| Road Distance<br>& Provider Category<sup>a</sup> |  |  | 48.7<br>(43.2-54.1) | -0.6% | 58.8<br>(54.1-63.5) | -2.5% | 37.5<br>(30.4-44.6) | 0.8% | 40.2<br>(32.5-47.9) | 0.2% | 20.2*<br>(16.0-24.4) | -11.8% | 16.5<br>(12.3-21.8) | -1.8% |
| Radius 5 km<br>& Provider Category<sup>a</sup> |  |  | 49.2<br>(43.7-54.7) | 0.4% | 59.4<br>(54.8-64.1) | -1.5% |  |  |  |  |  |  |  |  |
| Radius 10 km –<br>Unweighted<sup>b</sup> |  |  |  |  |  |  | 35.3*<br>(29.3-41.4) | -5.1% | 39.1<br>(32.0-46.2) | -2.5% | 18.8*<br>(14.9-22.7) | -17.9% | 15.7<br>(12.4-19.1) | -6.5% |
| Radius 10 km – Weighted<sup>c</sup> |  |  |  |  |  |  | 37.5<br>(30.6-44.4) | 0.8% | 39.8<br>(32.5-47.1) | -0.7% | 19.1*<br>(15.1-23.0) | -16.6% | 15.6<br>(12.1-19.1) | -7.1% |
| KDE - Single |  |  | 71.8*<br>(69.3-74.2) | 46.5% | 55*<br>(50.4-59.6) | -8.8% |  |  |  |  |  |  |  |  |
| KDE- Aggregate |  |  | 74.3*<br>(73.2-75.5) | 51.6% | 54.9*<br>(50.4-59.5) | -9.0% |  |  |  |  |  |  |  |  |
| **Ecological - Administrative** |  |  |  |  |  |  |  |  |  |  |  |  |  |  |
| Facility Catchment &<br>Provider Category<sup>a</sup> |  |  | 49.1<br>(43.6-54.6) | 0.2% | 59.8<br>(55.1-64.5) | -0.8% |  |  |  |  |  |  |  |  |
| Sub-District &<br>Provider Category<sup>a</sup> |  |  | 49.4<br>(43.9-54.9) | 0.8% | 57.9<br>(53.4-62.4) | -4.0% |  |  |  |  |  |  |  |  |
| District - Unweighted<sup>b</sup> | 4.7*<br>(21.4-31.7) | -52.5% |  |  |  |  | 34.9*<br>(29.0-40.8) | -6.2% | 39<br>(32.3-45.7) | -2.7% | 17.8*<br>(14.6-21.0) | -22.3% | 21*<br>(17.2-24.8) | 25.0% |
| District - Unweighted<sup>b</sup><br>& Provider Category<sup>a</sup> | 11.0<br>(3.8-18.2) | 11.8% |  |  |  |  | 37<br>(30.4-43.6) | -0.5% | 39.7<br>(32.7-46.7) | -1.0% | 20.3*<br>(15.8-24.8) | -11.4% | 17.4<br>(13.3-21.4) | 3.6% |
| District - Weighted<sup>c</sup> |  |  |  |  |  |  | 37.9<br>(31.3-44.4) | 1.9% | 40.7<br>(33.7-47.7) | 1.5% | 19.7*<br>(16.1-23.2) | -14.0% | 21.2*<br>(17.4-25.0) | 26.2% |
| District - Weighted<sup>c</sup><br>& Provider Category<sup>a</sup> |  |  |  |  |  |  | 38.8*<br>(31.9-45.7) | 4.3% | 40.8<br>(33.6-48.0) | 1.7% | 21.1*<br>(16.4-25.8) | -7.9% | 17.1<br>(13.1-21.2) | 1.8% |

<sup>a</sup> Ecological linking restricted to only providers within the category (type of outlet, managing authority, and facility level) reported by survey respondent
<sup>b</sup> Simple average of provider quality scores applied, not accounting for differentials in patient volume
<sup>c</sup> Provider quality scores weighted by provider utilization volume for relevant health area

#### Performance of measures of geographic proximity for ecological linking

Eight studies assessed the performance of geographic measures in assigning households or individuals to their reported source of healthcare. Four studies in sub-Saharan Africa compared the predicted source of care based on geographic proximity against the true source of care. They found straight-line and road distance performed similarly [64], high performance of shortest travel time method [65], and better performance of straight-line distance compared to road distance [31,37]. In the US, a more sophisticated approach (2-stage and 3-stage floating catchment area) performed better than alternatives methods in assigning households to their source of care [66]. Three studies in sub-Saharan Africa evaluated use of Theissen boundaries, a method of defining catchment boundaries based on the optimal distance between known providers, in assigning households to the catchment of facilities they utilized. The studies found high performance in some settings [67], but poorer performance related to the use of higher-order facilities [68] and influence of public transportation routes [69].

#### Statistical challenges

Most linking analyses that have generated effective coverage estimates by assigning individuals the quality score of the reported or linked source of care have derived estimates of uncertainty based on household sampling error and ignored any sampling error around provider data. However, two analyses used the Delta method (73) for estimating the variance of effective coverage estimates generated by multiplying service use and readiness [18,39]. These papers did not elaborate on why the method was chosen or compare the estimates to values generated using alternative approaches. No other papers in this analysis proposed or examined methods to derive measures of uncertainty for linked estimates.

## DISCUSSION

This review found a limited number of publications that explicitly addressed methodological issues related to linking household and provider datasets. A summary of key findings and needs for further research is presented in table 4 and discussed below.

**Table 4.**
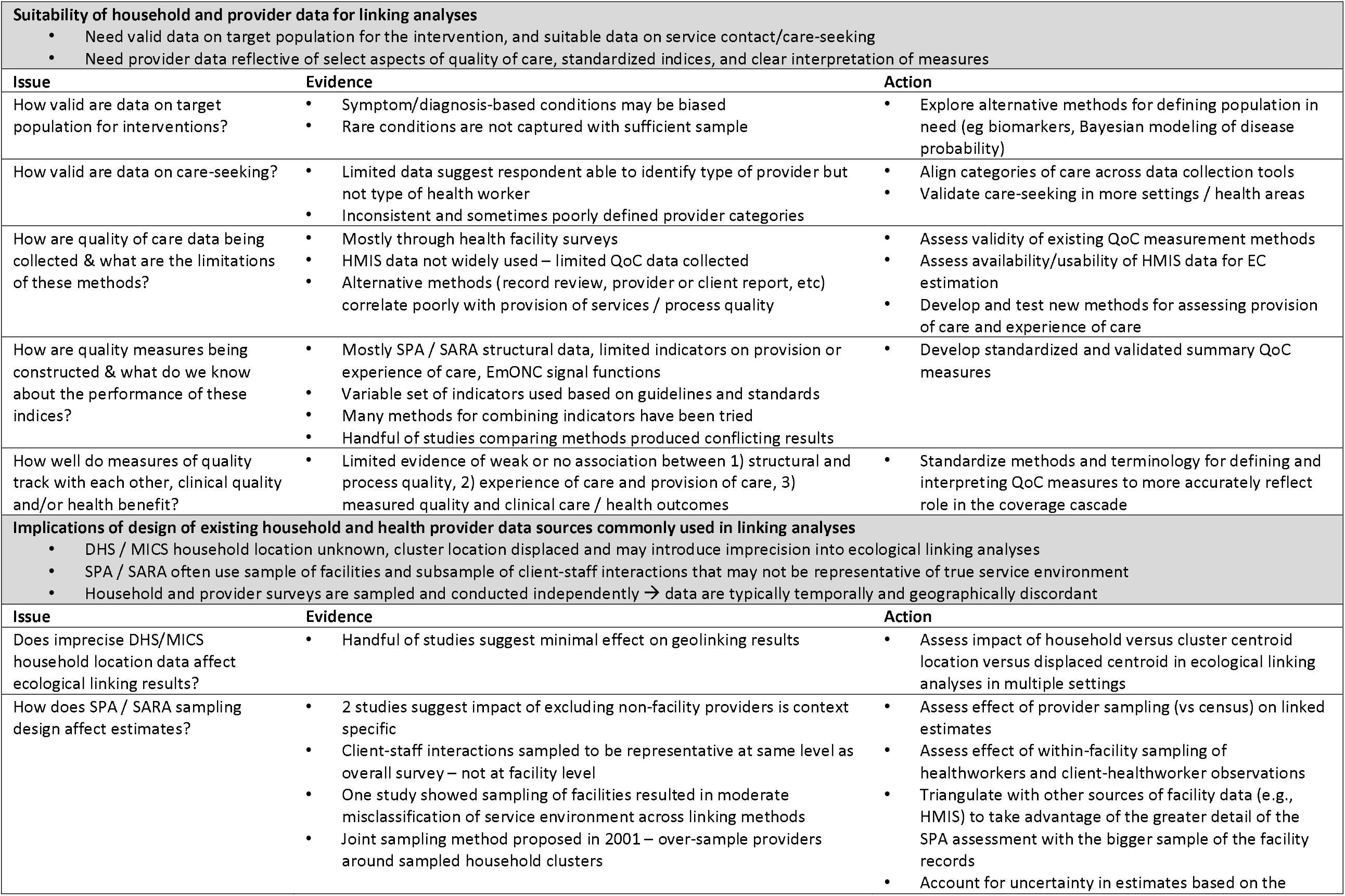
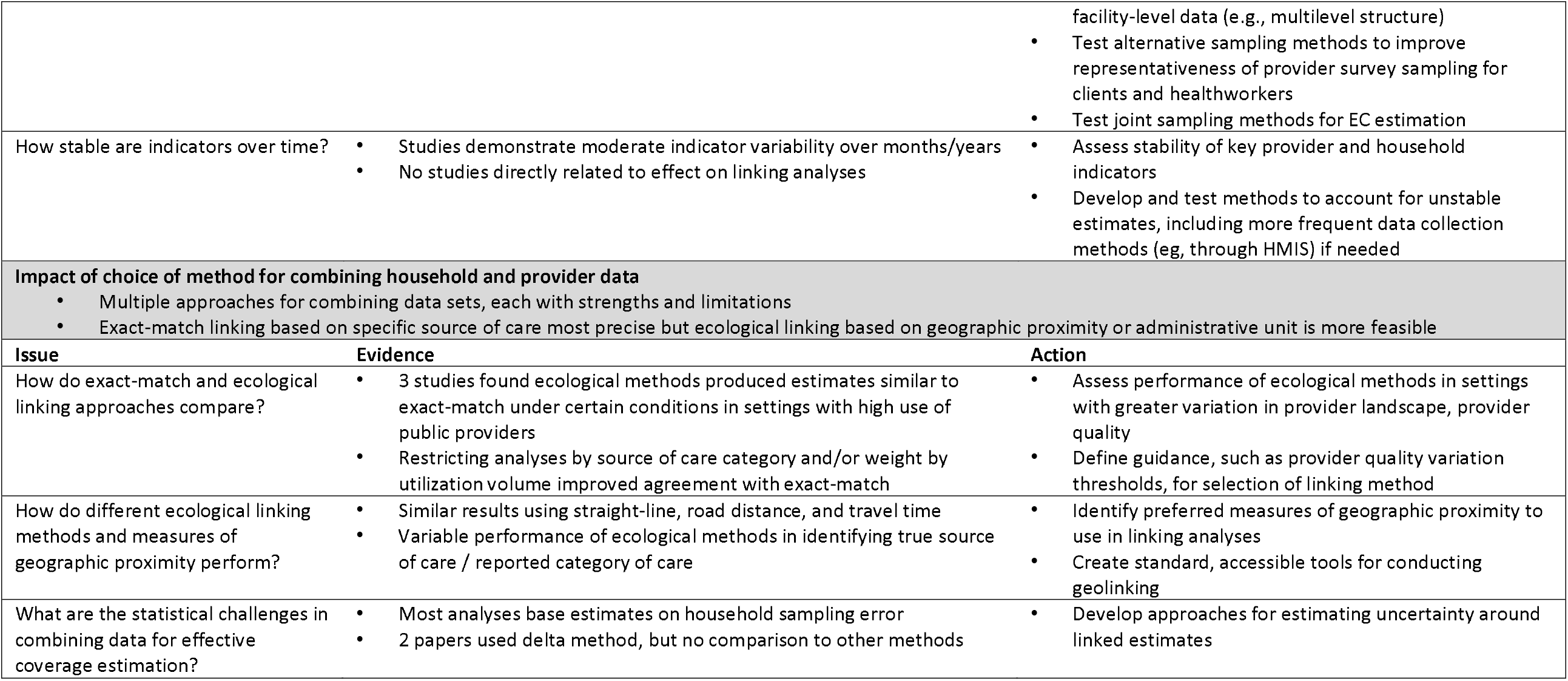
Summary of evidence related to methodological issues for linking analyses and related needs for future research

We identified a number of papers that critically assessed household and provider data needed for linking analyses. The limited existing data on respondent-reported care-seeking suggest respondents can identify sources of care if not individual healthcare worker cadre, but additional validation in various settings and service areas, such as postnatal care, would be informative. Further, it is essential to ensure that categorization of sources of care in household surveys align with the categories used in provider assessments to facilitate linking datasets. The validity of household survey data for estimating populations in need was more variable. While some populations in need can be clearly defined, others, particularly those requiring symptom-derived diagnoses based on respondent report, have demonstrated potential for bias. Additional work is needed to explore alternative methods for identifying populations in need in population-based data collection.

The content and construction of provider quality indices was highly variable across publications, but largely derived from facility surveys and was informed by international guidelines or recommendations. Methods for collecting provider quality have a number of limitations, and no single method perfectly encompasses all aspects of care [71]. The review found a lack of agreement between measures of quality derived through various means of collection. Overall, there was little empirical data supporting association between structural quality and process quality, and measures of quality and appropriate care or good health outcomes, although the number of reviewed studies was very limited. However, as articulated by Nguhiu and colleagues, there is need to consider quality indicators’ “intrinsic value as levers for management action” and application to policy decision-making in addition to their ability to capture or predict associated health gain [36]. Additional research is needed in the short term to develop and evaluate new quality indices utilizing existing data sources (e.g., facility surveys, HMIS, and medical records) with an aim of identifying a standardized approach for selecting, combining, and interpreting indicators that reflect aspects of provider quality necessary for delivering appropriate, respectful, and effective care. Longer term, substantial effort is needed to strengthen or adapt existing mechanisms or develop alternative methods for collecting provider quality indicators that can produce timely and informative estimates for tracking effective coverage of key interventions.

A handful of studies addressed the influence of the design of common data sources on linking analyses, including the impact of imprecise household GIS data, provider sampling frame and sampling design, and estimate stability. However, there was a lack of concrete evidence around the impact of these factors on linked effective coverage estimates. Explicitly evaluating the impact of imprecise household location, sampling design, and temporal gaps between measures within the context of effective coverage estimation would be informative. Mixed results on the inclusion of non-facility providers in provider assessments for effective coverage estimation emphasize the need to empirically assess the utilization and service quality of non-facility providers in a given setting prior to conducting a linking analysis. Although data related to impact on effective coverage estimation were limited, small samples of client-staff observations, sampling of facilities, and temporal gaps between household and provider data have the potential to bias estimates. The available data suggest that developing and testing alternative means of sampling health providers could improve the validity of linked estimates of effective coverage, including evaluating joint sampling approaches proposed by Measure Evaluation [59] or used by other data collection mechanisms such as PMA2020 and the India District Level Household and Facility Survey. No papers effectively addressed statistical concerns in combining household and provider data sources. There is a need to develop and test approaches for estimating uncertainty around linked effective coverage estimates.

The most consistent evidence found through the review was around methods for combining data sets. Three papers compared ecological and exact match linking and reported that ecological linking produced similar estimates to exact-match linking under certain conditions. The agreement between the three publications that compared exact-match and ecological linking is promising. Exact-match linking is considered the most precise method for generating linked estimates; however, ecological linking is often more feasible because it does not require information on exact source of care or data on all providers. The papers further point to the need to maintain data on type of provider from which care was sought or the relative volume of patients seen by providers in order to generate valid estimates of effective coverage. All three studies were conducted in rural sub-Saharan Africa in settings with high utilization of public sector health facilities; additional studies evaluating the performance of these methods in settings with a more diverse healthcare landscape would be informative. Other papers evaluated ecological linking approaches and found similar estimates of access to care or effective coverage using different approaches for assessing geographic proximity, although the ability of methods to capture true source of care was more variable. External to this review, additional data suggests individuals may not always utilize the closest source of care and may bypass providers in favor of providers offering better care [35,72]. These findings along with the analyses comparing exact-match and ecological linking approaches emphasize the need to carefully select methods for performing ecological linking and to control for true care-seeking behavior as much as possible by accounting for the type of provider from which care was sought or weighting by utilization in linking analyses.

Evidence across the review demonstrates the need for careful choice of methods, data sources, and indicators when conducting studies or analyses to link household and provider data for effective coverage estimation. An exploration of the precise effect of setting characteristics, such as variation in provider quality, on effective coverage estimates is needed to guide decision-making in the selection of linking methods. Once more of these issues have been evaluated, there will be a need for additional tools and guidance to facilitate use of these methods.

The review was limited by the diversity of terminology and fields related to the linking methodology. However, the use of multiple search strategies minimized likelihood of overlooking relevant publications. Additionally, the diversity of fields, approaches, and questions made it difficult to summarize the findings neatly, emphasizing the need for communication between researchers, more standard terminology, and, ideally, a cohesive research strategy going forward.

## CONCLUSIONS

Linking household and healthcare provider data is a promising approach that leverages existing data sources to generate more informative estimates of intervention coverage and care. These methods can potentially address limitations of both household and provider surveys to generate population-based estimates that reflect not only use of services, but also the content and quality of care received and the potential for health benefit. However, there is need for additional research to develop evidence-based, standardized best practices for these analyses. The most pressing priorities identified in this review are: 1) for those collecting data from health systems to explore methods to strengthen existing provider data collection mechanisms and promote temporal and geographic alignment with population-based measures, 2) for those collecting population-based data to address validity of self-reported intervention need and ensure indicators of access and utilization of care are measured to facilitate linking analyses, and 3) for those conducting linked analyses to standardize approaches for generating and interpreting effective coverage indicators, including sources of uncertainty, to ensure we are producing evidence that is harmonized, informative, and actionable for governments and valid for monitoring population health globally.

## Data Availability

As a review article, this article reports data from previously published studies.

## Author contributions

Conceived of the study design: EC MM. ICMJE criteria for authorship read and met: EC HL TM AA MM. Agree with manuscript and conclusions: EC HL TM AA MM. Conducted review: EC. Drafted paper: EC. All authors read, edited, and approved the manuscript.

## Data Sharing

As a review article, this article reports data from previously published studies.

## Ethics Approval

Ethical approval was not required as this review only included publicly available, published data.

## Patient and public involvement

Patients and/or the public were not involved in the design, or conduct, or reporting, or dissemination plans of this research.

## Patient consent for publication

Not required.

## Provenance and peer review

Not commissioned; externally peer reviewed.

## Footnotes

1 We define health providers as health care outlets such as health facilities, pharmacies, and community-based health workers

